# Tumor response-speed heterogeneity as a novel prognostic factor in patients with mCRC

**DOI:** 10.1101/2022.07.22.22277828

**Authors:** Junjia Liu, Xuefeng Wang, Ibrahim H. Sahin, Iman Imanirad, Seth I. Felder, Richard D. Kim, Hao Xie

## Abstract

**Purpose:** Differential tumor response to therapy is partially attributed to tumor heterogeneity. Additional efforts are needed to identify tumor heterogeneity parameters in response to therapy that are easily applicable in clinical practice. We aimed to describe tumor response-speed heterogeneity and evaluate its prognostic value in patients with metastatic colorectal cancer (mCRC).

**Patients and Methods:** Individual patient data from Amgen (NCT00364013) and Sanofi (NCT00305188; NCT00272051) trials were retrieved from Project Data Sphere. Patients in the Amgen 5-fluorouracil, leucovorin, oxaliplatin (FOLFOX) arm were used to establish response-speed heterogeneity. Its prognostic value was subsequently validated in the Sanofi FOLFOX arms and the Amgen panitumumab + FOLFOX arm. Kaplan-Meier method and Cox proportional hazards models were used for survival analyses.

**Results:** Patients with high response-speed heterogeneity in the Amgen FOLFOX cohort had significantly shorter (*P*<0.001) median progression-free survival (PFS) of 7.27 months (95%CI 6.12–7.96 months) and overall survival (OS) of 16.0 months (95%CI 13.8–18.2 months) than patients with low response-speed heterogeneity with median PFS of 9.41 months (95%CI 8.75– 10.89 months) and OS of 22.4 months (95%CI 20.1–26.7 months), respectively. Tumor response-speed heterogeneity was a poor prognostic factor of shorter PFS (HR 4.17, 95%CI 2.49–6.99, *P*<0.001) and shorter OS (HR 2.57, 95%CI 1.64–4.01, *P*<0.001), after adjustment for other common prognostic factors. Comparable findings were found in the external validation cohorts.

**Conclusion:** Tumor response-speed heterogeneity to first-line chemotherapy was a novel prognostic factor associated with early disease progression and shorter survival in patients with mCRC.

**Implications for Practice:** Routine clinical decision making heavily relies on radiographic assessment of disease response to therapy. For patients with heterogeneous tumors, the degree and kinetics of individual tumor response to the same therapy can sometimes be vastly different. We explored a novel quantitative parameter to describe response-speed heterogeneity by utilizing individual patient data from previous clinical trials. This parameter was an independent prognostic factor associated with early disease progression and shorter survival. Complementary to existing molecular and radiographic tumor heterogeneity parameters, it may help practicing oncologists describe tumor response disparity and serve as a new prognostic factor for patients with mCRC.

## Introduction

Colorectal cancer is the third leading cause of cancer mortality in the United States.^[1]^ The five-year survival rate of patients with metastatic colorectal cancer (mCRC) is less than 15%.^[2]^ Genetic profiling of tumor samples from biopsy has become the cornerstone for the management of mCRC.^[3]^ Nevertheless, therapies matched with genetic profiling along with chemotherapy remain to provide limited clinical benefits, at least partially due to both inter- and intra-tumor heterogeneity.^[4]^

Tumor heterogeneity among metastatic sites develop from subclones of a primary lesion and with accumulation of additional molecular alterations throughout the disease course.^[5]^ Studies on tumor heterogeneity from different perspectives have been undertaken. For example, tumor heterogeneity can be assessed by molecular profiling with phylogenetic analyses of tumor samples at different metastatic sites and within individual lesions.^[6,7]^ However, implementation of this approach to assess tumor heterogeneity requires multiple and frequent invasive biopsy procedures, thus not clinically practical. In contrast, longitudinal radiographic assessments of tumor heterogeneity are readily available by analyzing features of image texture such as gray-level intensity and pixel positions within a CT, MRI or PET image. The heterogeneity of these features may reflect the underlying heterogeneity of tumor biology.^[8-10]^ Similar to molecular assessment of spatial and temporal tumor heterogeneity, this approach has not been widely implemented in clinical practice either, likely due to the requirement of high computational capability and implementation of complex machine learning algorithms in addition to further validation of its clinical benefits.^[11]^

Routine clinical decision making heavily relies on radiographic assessment of disease response to therapy. This is largely based on the change of the number and size of the lesions per Response Evaluation Criteria in Solid Tumors (RECIST) criteria.^[12]^ For most of the patients with heterogeneous tumors, the degree and kinetics of individual tumor response to the same therapy can sometimes be vastly different. This phenomenon has been widely acknowledged clinically and associated with poor outcome in some previous studies in other cancer types.^[13-16]^ But tumor response-speed heterogeneity has not been quantitatively defined and adequately studied using high quality clinical trials data in patients with mCRC. A recent study constructed a tumor heterogeneity parameter using Gower distance based on a complex mathematical formula with five variables.^[17]^ Gower distance with variables extracted from radiographic response data even beyond disease progression towards the end of the treatment course has nicely correlated with patients, outcome from different treatments. Additional efforts are still needed to identify tumor heterogeneity parameters that are readily calculated and easily applicable in clinical practice for patients with mCRC.

In this study, we aimed to establish a parameter to describe tumor response-speed heterogeneity and evaluate its prognostic value in patients with mCRC.

## Patients and Methods

### Patients

De-identified data of individual patients with previously untreated mCRC from Amgen (NCT00364013) and Sanofi (NCT00305188; NCT00272051) trials were retrieved from the Project Data Sphere.^[18]^ Analysis of these de-identified, publicly available data in the Project Data Sphere is in compliance with Health Insurance Portability and Accountability Act and exempted from review by the institutional review board.

NCT00364013 was a randomized phase 3 trial to assess the efficacy of panitumumab in addition to 5-fluorouracil, leucovorin, oxaliplatin (FOLFOX) as first-line therapy for patients with mCRC (two arms: panitumumab + FOLFOX4; FOLFOX4 alone). Primary endpoint was progression-free survival (PFS). Patients received treatment every 2 weeks until disease progression based on radiographic assessment every 8 weeks or until unacceptable toxicity. Patients were subsequently followed every 3 months until 30 months after the last patient was randomized.^[18, 19]^

NCT00305188 was a randomized phase 3 trial to study the efficacy of xaliproden in preventing the neurotoxicity of oxaliplatin in patients with previously untreated mCRC (two arms: xaliproden + mFOLFOX6; placebo + mFOLFOX6). Clinical evaluation of peripheral sensory neuropathy was the primary endpoint. PFS and overall survival (OS) were secondary endpoints. Xaliproden or placebo was administered daily. Chemotherapy was given every 2 weeks until disease progression. Tumors were evaluated radiographically every 8 weeks. Patients were followed until disease progression, or 13 months after the first cycle of chemotherapy, whichever came first.^[18, 20]^

Similarly, NCT00272051 was a randomized phase 3 trial to study the efficacy of xaliproden in reducing the neurotoxicity of oxaliplatin in patients with previously untreated mCRC (two arms: xaliproden + FOLFOX4; placebo + FOLFOX4). Primary endpoints were clinical evaluation of peripheral sensory neuropathy and response rate. PFS and OS were secondary endpoints. Xaliproden or placebo was administered daily. Chemotherapy was administered every 2 weeks up to 1 year. Tumor was evaluated radiographically every 8 weeks. Patients were followed until disease progression or for 13 months after the first dose of study drug, whichever came first.^[18, 21]^

In this study, we used patients in the FOLFOX arm of the Amgen study “Amgen (FOLFOX)” to establish response-speed heterogeneity. We then used patients in the FOLFOX arms of the Sanofi studies “Sanofi (FOLFOX)” and the panitumumab + FOLFOX arm of the Amgen study “Amgen (panitumumab + FOLFOX)” as the validation cohorts (**Supplemental Figure S1**). A minimum of two baseline target lesions are required to calculate the heterogeneity parameters. Thus, patients with fewer metastatic target lesions were excluded from the analysis. Patients with erroneous data records or missing critical information (e.g., baseline radiographic measurements of target lesions were missing.) were also excluded.

Baseline demographic, clinical, and radiographic variables used for analyses included those that were directly available from the data files or computed from the available data at the Project Data Sphere. PFS was defined as from randomization to disease progression per modified RECIST criteria. OS was defined as the time from randomization to death. Sum of tumor size at the baseline and each treatment visit was provided in the Amgen data files, and was calculated by summing the size of each target lesion at each visit in the Sanofi trials. Data from local radiographic assessments (e.g., tumor sizes, locations, etc.) were used in our study for patients from the Amgen trial.

### Response-speed heterogeneity

Response-speed heterogeneity (H_r_) reflects the variability between target lesions, individual response speed (r), which was defined as the average relative-size change speed from baseline to each lesion,s best response (lowest lesion size in measurement).

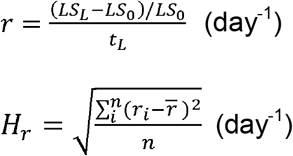

Where LS_o_ is the baseline lesion size (LS) of a target lesion, LS_L_ is its lowest LS after at least 4 weeks of treatment and before the time of disease progression, and t_L_ is the time from the start of treatment to LS_L_, 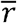 is the mean of r within a patient, and n is the number of target lesions.

Overall response speed (R) was defined as the average relative-size change speed of sum size (SS) of lesion measurement from baseline to the best response (lowest SS), reflecting the overall response.

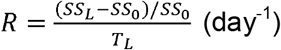

Where SS_o_ is the baseline SS of target lesions in a patient, SS_L_ is its lowest SS after at least 4 weeks of treatment and before the time of disease progression, and T_L_ is the time from the start of treatment to SS_L_.

Baseline size heterogeneity (H_bl_) was defined to reflect the variability between baseline LS of target lesions in a patient.

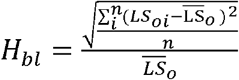

We described how individual and sum lesion sizes change with time upon exposure to the treatment in representative patients in **Supplemental Figure S2**.

### Statistical analysis

Continuous variables were summarized as median, interquartile ranges (IQR), and compared using the Wilcoxon rank-sum test or Kruskal-Wallis test. Categorical variables were summarized as frequency counts, percentages and compared using the Fisher exact test. Univariate and multivariable Cox proportional hazards models were used to evaluate prognostic factors. Clinically relevant variables were included in univariate and multivariable models. Results from all Cox proportional hazards models were presented as hazard ratios (HR), 95% confidence interval (CI) for the HR, and corresponding *P*-values. Time-to-event data were summarized using Kaplan-Meier method with median of the response-speed heterogeneity as the cutoff for dichotomization, and compared using the log-rank test. All tests were two-sided and performed in R (version 3.6, R Foundation for Statistical Computing, Vienna, Austria) or in Python (version 3.8). A *P*-value <0.05 was considered statistically significant in this study.

## Results

### Patients

This study included 935 patients from the Amgen trial and 756 patients from the two Sanofi trials combined. Patients, baseline demographic, clinical, and tumor radiographic characteristics were summarized in **Table 1**. The Amgen trial had higher percentages of white/Caucasian patients, patients with higher Eastern Cooperative Oncology Group (ECOG) performance score, patients with higher number of metastatic sites, patients with liver metastasis, and patients with lymph node metastasis, lower carcinoembryonic antigen (CEA), and longer follow-up time, compared to the Sanofi trials.

**Table 1.**
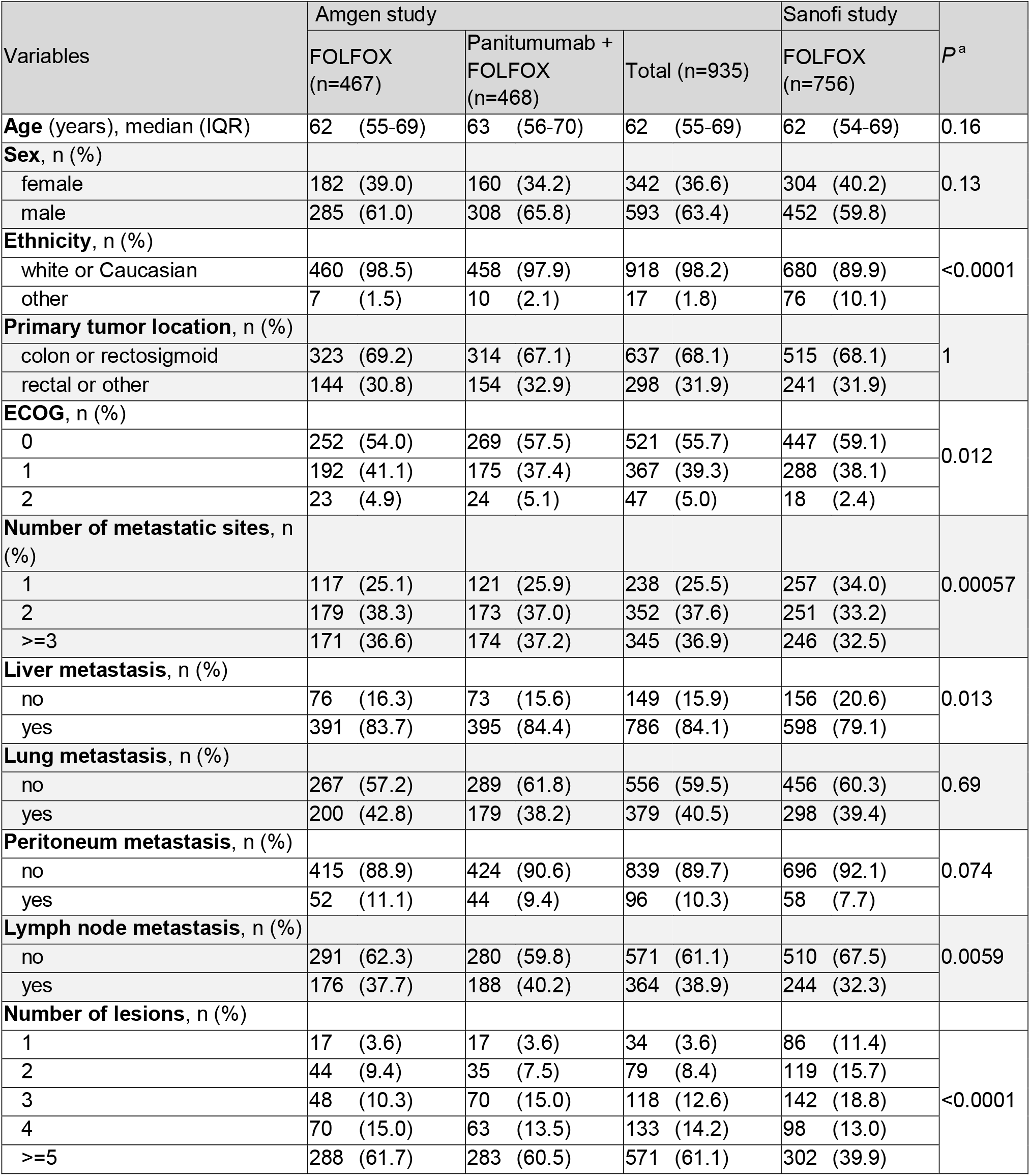
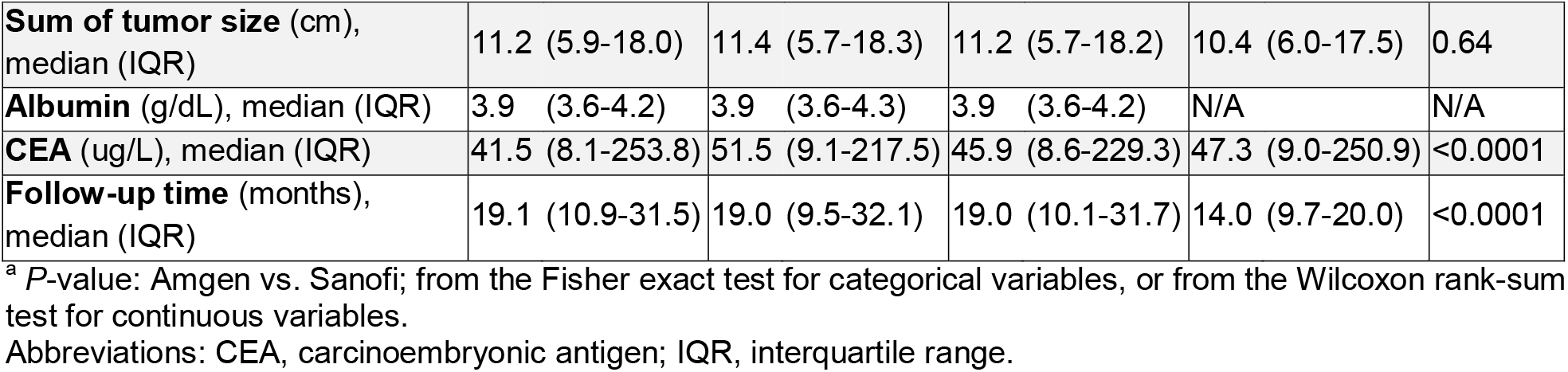
Baseline characteristics of patients in the trials.

| Variables | Amgen study |  |  | Sanofi study |  | P<sup>a</sup> |
| --- | --- | --- | --- | --- | --- | --- |
|  | FOLFOX<br>(n=467) | Panitumumab +<br>FOLFOX<br>(n=468) | Total (n=935) | FOLFOX<br>(n=756) |  |  |
| **Age** (years), median (IQR) | 62 (55-69) | 63 (56-70) | 62 (55-69) | 62 (54-69) |  | 0.16 |
| **Sex**, n (%) |  |  |  |  |  |  |
| female | 182 (39.0) | 160 (34.2) | 342 (36.6) | 304 (40.2) |  | 0.13 |
| male | 285 (61.0) | 308 (65.8) | 593 (63.4) | 452 (59.8) |  |  |
| **Ethnicity**, n (%) |  |  |  |  |  |  |
| white or Caucasian | 460 (98.5) | 458 (97.9) | 918 (98.2) | 680 (89.9) |  | <0.0001 |
| other | 7 (1.5) | 10 (2.1) | 17 (1.8) | 76 (10.1) |  |  |
| **Primary tumor location**, n (%) |  |  |  |  |  |  |
| colon or rectosigmoid | 323 (69.2) | 314 (67.1) | 637 (68.1) | 515 (68.1) |  | 1 |
| rectal or other | 144 (30.8) | 154 (32.9) | 298 (31.9) | 241 (31.9) |  |  |
| **ECOG**, n (%) |  |  |  |  |  |  |
| 0 | 252 (54.0) | 269 (57.5) | 521 (55.7) | 447 (59.1) |  | 0.012 |
| 1 | 192 (41.1) | 175 (37.4) | 367 (39.3) | 288 (38.1) |  |  |
| 2 | 23 (4.9) | 24 (5.1) | 47 (5.0) | 18 (2.4) |  |  |
| **Number of metastatic sites**, n (%) |  |  |  |  |  |  |
| 1 | 117 (25.1) | 121 (25.9) | 238 (25.5) | 257 (34.0) |  | 0.00057 |
| 2 | 179 (38.3) | 173 (37.0) | 352 (37.6) | 251 (33.2) |  |  |
| >=3 | 171 (36.6) | 174 (37.2) | 345 (36.9) | 246 (32.5) |  |  |
| **Liver metastasis**, n (%) |  |  |  |  |  |  |
| no | 76 (16.3) | 73 (15.6) | 149 (15.9) | 156 (20.6) |  | 0.013 |
| yes | 391 (83.7) | 395 (84.4) | 786 (84.1) | 598 (79.1) |  |  |
| **Lung metastasis**, n (%) |  |  |  |  |  |  |
| no | 267 (57.2) | 289 (61.8) | 556 (59.5) | 456 (60.3) |  | 0.69 |
| yes | 200 (42.8) | 179 (38.2) | 379 (40.5) | 298 (39.4) |  |  |
| **Peritoneum metastasis**, n (%) |  |  |  |  |  |  |
| no | 415 (88.9) | 424 (90.6) | 839 (89.7) | 696 (92.1) |  | 0.074 |
| yes | 52 (11.1) | 44 (9.4) | 96 (10.3) | 58 (7.7) |  |  |
| **Lymph node metastasis**, n (%) |  |  |  |  |  |  |
| no | 291 (62.3) | 280 (59.8) | 571 (61.1) | 510 (67.5) |  | 0.0059 |
| yes | 176 (37.7) | 188 (40.2) | 364 (38.9) | 244 (32.3) |  |  |
| **Number of lesions**, n (%) |  |  |  |  |  |  |
| 1 | 17 (3.6) | 17 (3.6) | 34 (3.6) | 86 (11.4) |  | <0.0001 |
| 2 | 44 (9.4) | 35 (7.5) | 79 (8.4) | 119 (15.7) |  |  |
| 3 | 48 (10.3) | 70 (15.0) | 118 (12.6) | 142 (18.8) |  |  |
| 4 | 70 (15.0) | 63 (13.5) | 133 (14.2) | 98 (13.0) |  |  |
| >=5 | 288 (61.7) | 283 (60.5) | 571 (61.1) | 302 (39.9) |  |  |

|  |  |  |  |  |  |
| --- | --- | --- | --- | --- | --- |
| **Sum of tumor size** (cm), median (IQR) | 11.2 (5.9-18.0) | 11.4 (5.7-18.3) | 11.2 (5.7-18.2) | 10.4 (6.0-17.5) | 0.64 |
| **Albumin** (g/dL), median (IQR) | 3.9 (3.6-4.2) | 3.9 (3.6-4.3) | 3.9 (3.6-4.2) | N/A | N/A |
| **CEA** (ug/L), median (IQR) | 41.5 (8.1-253.8) | 51.5 (9.1-217.5) | 45.9 (8.6-229.3) | 47.3 (9.0-250.9) | <0.0001 |
| **Follow-up time** (months), median (IQR) | 19.1 (10.9-31.5) | 19.0 (9.5-32.1) | 19.0 (10.1-31.7) | 14.0 (9.7-20.0) | <0.0001 |

<sup>a</sup> *P*-value: Amgen vs. Sanofi; from the Fisher exact test for categorical variables, or from the Wilcoxon rank-sum test for continuous variables.
Abbreviations: CEA, carcinoembryonic antigen; IQR, interquartile range.

### Response-speed heterogeneity

The median response-speed heterogeneity was 1.19 × 10^−3^ day^-1^ (IQR 0.60 × 10^−3^ – 2.47 × 10^−3^ day^-1^) in the Amgen (FOLFOX) cohort, 1.58 × 10^−3^ day^-1^ (IQR 0.66 × 10^−3^ – 3.23 × 10^−3^ day^-1^) in the Sanofi (FOLFOX) cohort, and 1.24 × 10^−3^ day^-1^ (IQR 0.56 × 10^−3^ – 2.50 × 10^−3^ day^-1^) in the Amgen (panitumumab + FOLFOX) cohort. The response-speed heterogeneity was associated with higher number of metastatic sites, presence of lymph node metastasis, higher number of metastatic lesions, and higher sum size of target lesions at baseline, and progressive disease at first radiographic evaluation in the Amgen (FOLFOX) cohort (**Figure 1**; **Supplemental Figure S3**).

**Figure 1.**
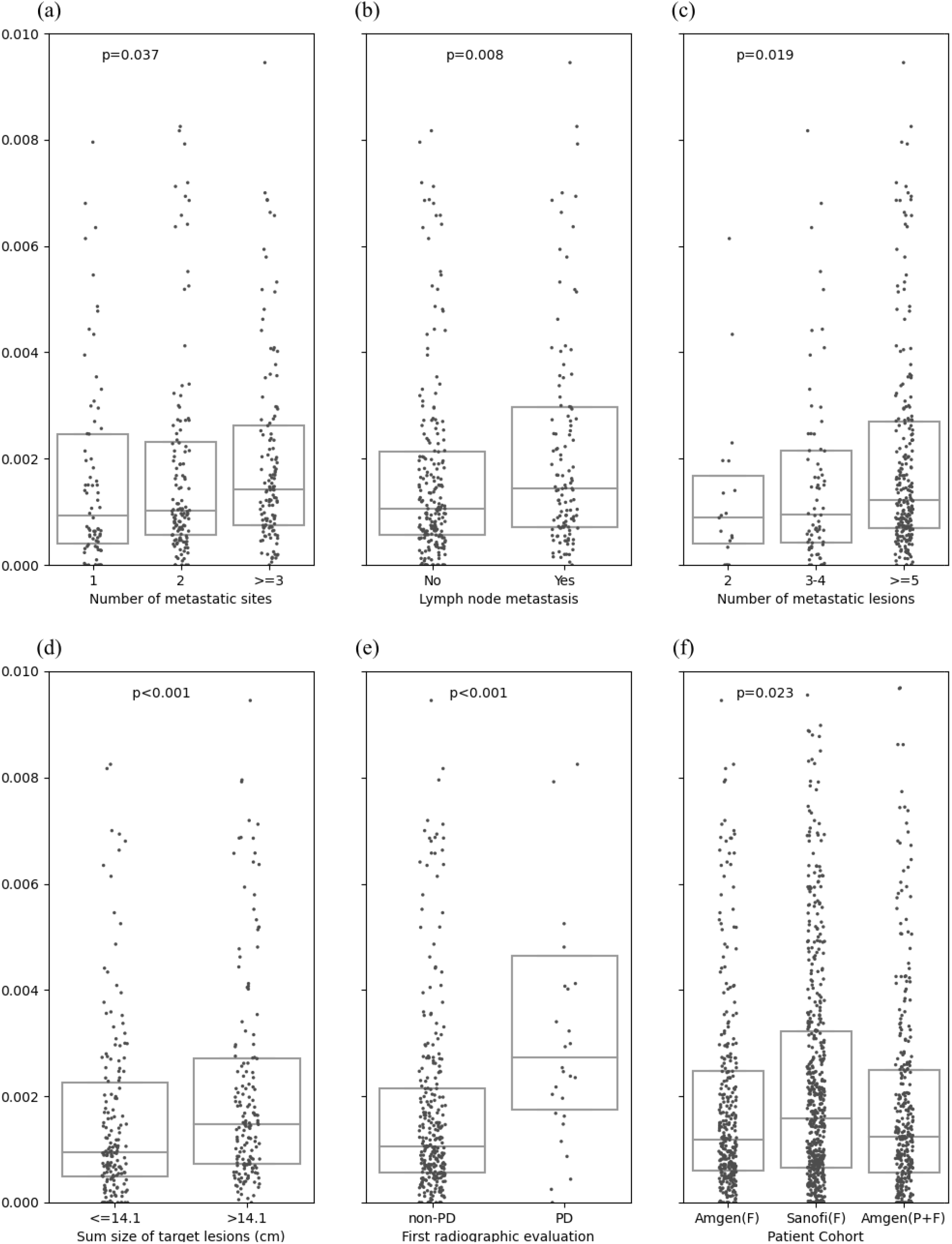
Distributions of response-speed heterogeneity stratified by baseline (a) number of metastatic sites, (b) presence of lymph node metastasis, (c) number of metastatic lesions, (d) sum size of target lesions, and (e) first radiographic evaluation in the Amgen (FOLFOX) cohort; and (f) patients in the Amgen (FOLFOX), Sanofi (FOLFOX), and Amgen (panitumumab + FOLFOX) cohorts. *P*-value from Wilcoxon rank-sum test for comparing two groups, or from Kruskal-Wallis test for comparing three or more groups.

### Association with PFS

Patients in the Amgen (FOLFOX) cohort were stratified into response-speed heterogeneity high versus low groups. Patients with high response-speed heterogeneity had a significantly shorter (*P=*0.0004) median PFS of 7.27 months (95% CI 6.12–7.96 months) than patients with low response-speed heterogeneity with median PFS of 9.41 months (95% CI 8.75– 10.89 months) (**Figure 2a**). Univariate survival analysis of the Amgen (FOLFOX) cohort showed that higher response-speed heterogeneity among other traditional prognostic factors was significantly associated with shorter PFS with HR 2.99 (95% CI 1.85–4.86, *P<*0.0001) (**Table 2**). This observation holds true in multivariable survival analysis where higher response-speed heterogeneity was independently associated with shorter PFS with HR 4.17 (95% CI 2.49–6.99, P<0.0001) after adjustment for other common prognostic factors including baseline sum size of target lesions, number of metastatic lesions, ECOG, presence of liver metastasis, presence of lung metastasis, albumin, and response speed. A sensitivity analysis to evaluate the association of response-speed heterogeneity with PFS in the Amgen (FOLFOX) cohort using patients with only ≥2 target lesions in the liver demonstrated comparable results (**Supplemental Table S1**).

**Figure 2.**
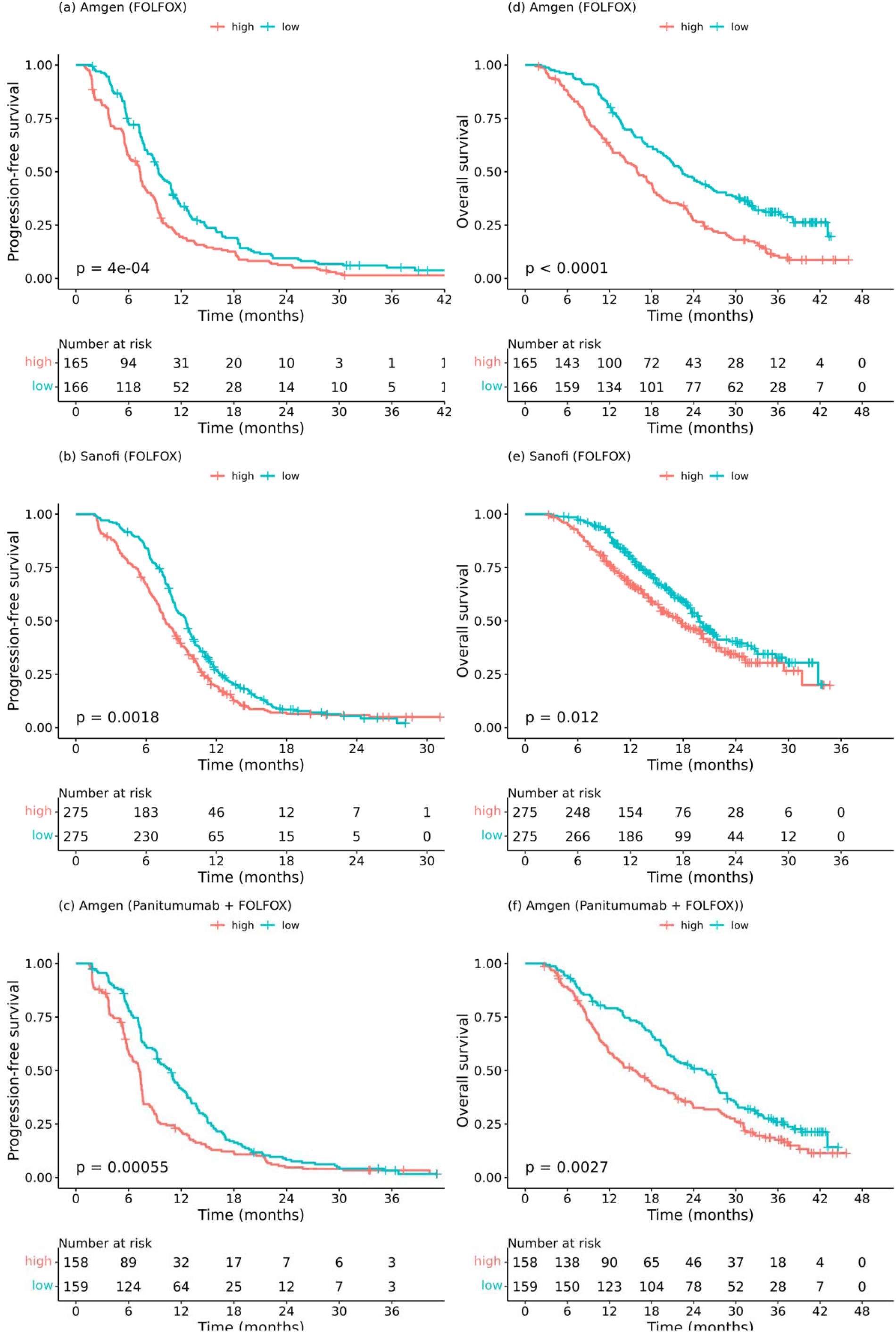
Survival analyses stratified by high and low groups (median as the cutoff) of response-speed heterogeneity. Kaplan-Meier curves of PFS in (a) the Amgen (FOLFOX) cohort, (b) the Sanofi (FOLFOX) cohort, and (c) the Amgen (panitumumab + FOLFOX) cohort, and OS in (d) the Amgen (FOLFOX) cohort, (e) the Sanofi (FOLFOX) cohort, and (f) the Amgen (panitumumab + FOLFOX) cohort.

**Table 2.**
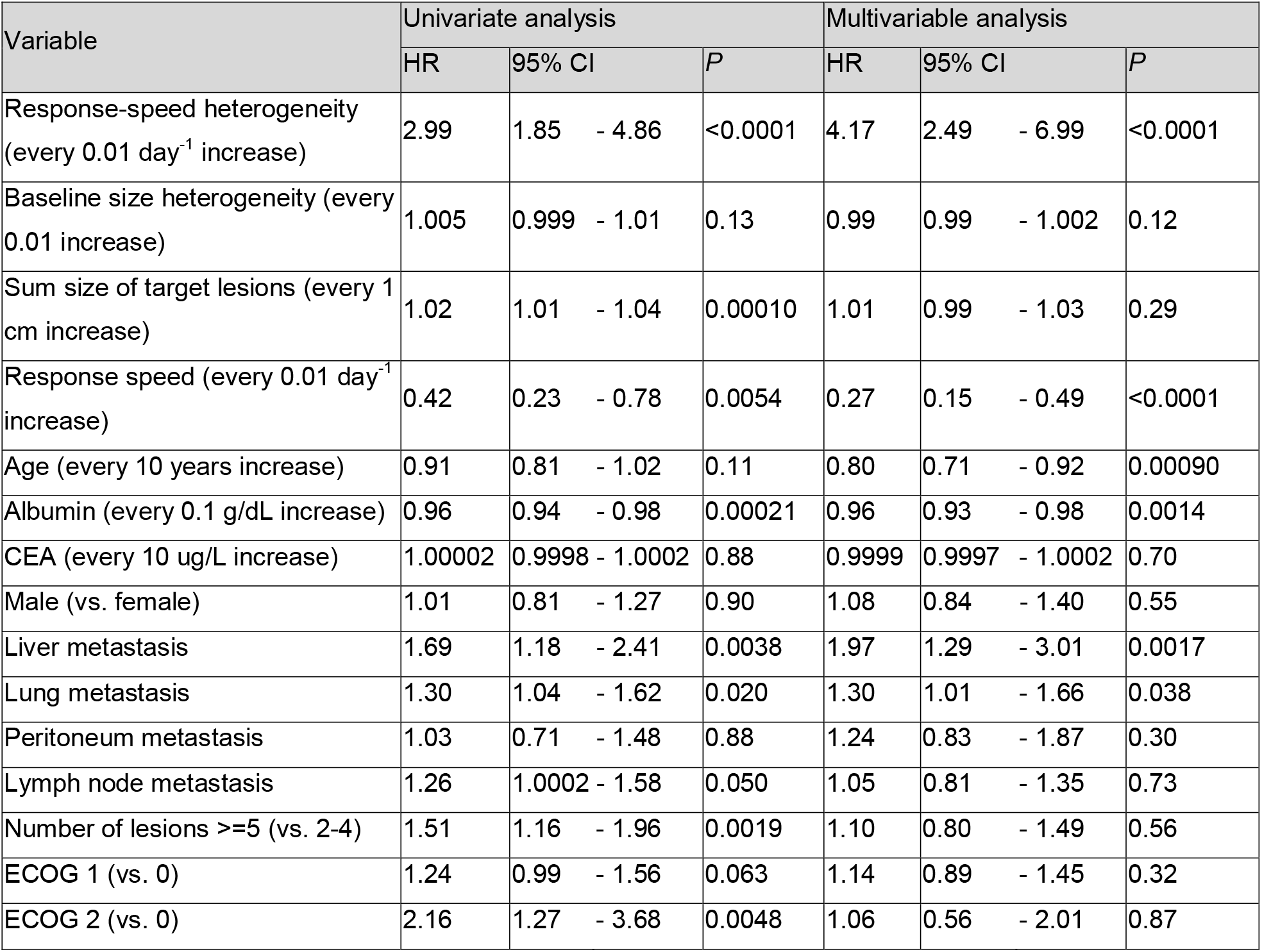
Univariate and multivariable analyses of PFS for the Amgen (FOLFOX) cohort.

| Variable | Univariate analysis |  |  | Multivariable analysis |  |  |
| --- | --- | --- | --- | --- | --- | --- |
|  | HR | 95% CI | *P* | HR | 95% CI | *P* |
| Response-speed heterogeneity (every 0.01 day<sup>-1</sup> increase) | 2.99 | 1.85 - 4.86 | <0.0001 | 4.17 | 2.49 - 6.99 | <0.0001 |
| Baseline size heterogeneity (every 0.01 increase) | 1.005 | 0.999 - 1.01 | 0.13 | 0.99 | 0.99 - 1.002 | 0.12 |
| Sum size of target lesions (every 1 cm increase) | 1.02 | 1.01 - 1.04 | 0.00010 | 1.01 | 0.99 - 1.03 | 0.29 |
| Response speed (every 0.01 day<sup>-1</sup> increase) | 0.42 | 0.23 - 0.78 | 0.0054 | 0.27 | 0.15 - 0.49 | <0.0001 |
| Age (every 10 years increase) | 0.91 | 0.81 - 1.02 | 0.11 | 0.80 | 0.71 - 0.92 | 0.00090 |
| Albumin (every 0.1 g/dL increase) | 0.96 | 0.94 - 0.98 | 0.00021 | 0.96 | 0.93 - 0.98 | 0.0014 |
| CEA (every 10 ug/L increase) | 1.00002 | 0.9998 - 1.0002 | 0.88 | 0.9999 | 0.9997 - 1.0002 | 0.70 |
| Male (vs. female) | 1.01 | 0.81 - 1.27 | 0.90 | 1.08 | 0.84 - 1.40 | 0.55 |
| Liver metastasis | 1.69 | 1.18 - 2.41 | 0.0038 | 1.97 | 1.29 - 3.01 | 0.0017 |
| Lung metastasis | 1.30 | 1.04 - 1.62 | 0.020 | 1.30 | 1.01 - 1.66 | 0.038 |
| Peritoneum metastasis | 1.03 | 0.71 - 1.48 | 0.88 | 1.24 | 0.83 - 1.87 | 0.30 |
| Lymph node metastasis | 1.26 | 1.0002 - 1.58 | 0.050 | 1.05 | 0.81 - 1.35 | 0.73 |
| Number of lesions ≥5 (vs. 2-4) | 1.51 | 1.16 - 1.96 | 0.0019 | 1.10 | 0.80 - 1.49 | 0.56 |
| ECOG 1 (vs. 0) | 1.24 | 0.99 - 1.56 | 0.063 | 1.14 | 0.89 - 1.45 | 0.32 |
| ECOG 2 (vs. 0) | 2.16 | 1.27 - 3.68 | 0.0048 | 1.06 | 0.56 - 2.01 | 0.87 |

These findings were validated in the external cohort, Sanofi (FOLFOX). Patients with high response-speed heterogeneity had a significantly shorter (P=0.0018) median PFS of 7.63 months (95% CI 7.14–8.45 months) than patients with low response-speed heterogeneity with median PFS of 9.41 months (95% CI 8.59–9.84 months) (**Figure 2b**). The association of higher response-speed heterogeneity with shorter PFS was confirmed in univariate (**Supplemental Table S2**) and multivariable survival analysis with HR of 1.67 (95% CI 1.10–2.54, *P=*0.016) after adjustment for other prognostic factors (**Table 3**).

**Table 3.** Multivariable analyses of PFS for the Sanofi (FOLFOX) and Amgen (panitumumab + FOLFOX) cohorts.

| Variable | Sanofi (FOLFOX)<br>Multivariable analysis |  |  | Amgen (panitumumab + FOLFOX)<br>Multivariable analysis |  |  |
| --- | --- | --- | --- | --- | --- | --- |
|  | HR | 95% CI | P | HR | 95% CI | P |
| Response-speed heterogeneity (every 0.01 day<sup>-1</sup> increase) | 1.67 | 1.10 - 2.54 | 0.016 | 2.37 | 1.35 - 4.14 | 0.0025 |
| Baseline size heterogeneity (every 0.01 increase) | 0.9997 | 0.995 - 1.004 | 0.90 | 0.999 | 0.99 - 1.01 | 0.80 |
| Sum size of target lesions (every 1 cm increase) | 1.02 | 1.004 - 1.03 | 0.012 | 0.99 | 0.98 - 1.01 | 0.48 |
| Response speed (every 0.01 day<sup>-1</sup> increase) | 0.38 | 0.24 - 0.60 | <0.0001 | 0.53 | 0.31 - 0.93 | 0.026 |
| Age (every 10 years increase) | 0.92 | 0.85 - 1.004 | 0.063 | 1.05 | 0.91 - 1.21 | 0.47 |
| Albumin (every 0.1 g/dL increase) | N/A | N/A | N/A | 0.98 | 0.95 - 1.003 | 0.089 |
| CEA (every 10 ug/L increase) | 0.9996 | 0.999 - 1.0004 | 0.35 | 1.0002 | 0.9998 - 1.001 | 0.28 |
| Male (vs. female) | 0.86 | 0.71 - 1.05 | 0.14 | 0.88 | 0.68 - 1.13 | 0.31 |
| Liver metastasis | 1.41 | 1.05 - 1.88 | 0.022 | 0.74 | 0.48 - 1.13 | 0.17 |
| Lung metastasis | 1.47 | 1.21 - 1.78 | 0.00011 | 0.90 | 0.70 - 1.16 | 0.41 |
| Peritoneum metastasis | 1.38 | 0.94 - 2.03 | 0.097 | 1.17 | 0.76 - 1.78 | 0.48 |
| Lymph node metastasis | 1.02 | 0.83 - 1.26 | 0.83 | 0.94 | 0.73 - 1.20 | 0.62 |
| Number of lesions >=5 (vs. 2-4) | 0.91 | 0.73 - 1.14 | 0.42 | 1.22 | 0.89 - 1.68 | 0.22 |
| ECOG 1 (vs. 0) | 1.06 | 0.87 - 1.29 | 0.54 | 1.31 | 0.99 - 1.72 | 0.055 |
| ECOG 2 (vs. 0) | 1.36 | 0.69 - 2.65 | 0.37 | 2.39 | 1.28 - 4.45 | 0.0062 |

These findings were also validated in a cohort of patients Amgen (panitumumab + FOLFOX) with similar baseline characteristics but receipt of a different treatment. Patients with high response-speed heterogeneity had a significantly shorter (*P*=0.00055) median PFS of 7.24 months (95% CI 6.18–7.47 months) than patients with low response-speed heterogeneity with median PFS of 10.82 months (95% CI 9.24–12.01 months) (**Figure 2c)**. The association of higher response-speed heterogeneity with shorter PFS was confirmed in univariate (**Supplemental Table S3**) and multivariable survival analysis with HR of 2.37 (95% CI 1.35– 4.14, *P=*0.0025) after adjustment for other prognostic factors (**Table 3**).

### Association with OS

In the Amgen (FOLFOX) cohort, patients with high response-speed heterogeneity had a significantly shorter (*P<*0.0001) median OS of 16.0 months (95% CI 13.8–18.2 months) than patients with low response-speed heterogeneity with median OS of 22.4 months (95% CI 20.1– 26.7 months). (**Figure 2d**). Multivariable survival analysis showed that higher response-speed heterogeneity was independently associated with shorter OS with HR of 2.57 (95% CI 1.64– 4.01, *P<*0.0001) after adjustment for similar prognostic factors mentioned above (**Supplemental Table S4**).

In the Sanofi (FOLFOX) cohort, patients with high response-speed heterogeneity had a significant shorter (*P=*0.012) median OS of 17.5 months (95% CI 15.4–20.3 months) than patients with low response-speed heterogeneity with median OS of 19.8 months (95% CI 18.7– 21.9 months) (**Figure 2e**). Multivariable survival analysis showed that higher response-speed heterogeneity had a trend of shorter OS with HR of 1.55 (95% CI 0.91–2.66, *P=*0.11) after adjustment for other prognostic factors (**Supplemental Table S5**).

In the Amgen (panitumumab + FOLFOX) cohort, patients with high response-speed heterogeneity had a significant shorter (*P=*0.0027) median OS of 15.8 months (95% CI 12.4– 19.8 months) than patients with low response-speed heterogeneity with median OS of 25.8 months (95% CI 20.3–27.5 months) (**Figure 2f**). Multivariable survival analysis showed that higher response-speed heterogeneity was independently associated with shorter OS with HR of 1.88 (95% CI 1.04–3.39, *P=*0.037) after adjustment for other prognostic factors (**Supplemental Table S6**).

## Discussion

Our study utilizing individual patient data from previous clinical trials established a novel parameter to quantify response-speed heterogeneity among individual target lesions in patients with newly diagnosed mCRC receiving first-line therapy. The patient populations used to develop the response-speed heterogeneity parameter and subjected for subsequent validations were largely representative of the real-world patient population. This statement was supported by comparable baseline demographic, clinical characteristics and common prognostic factors found in patients with mCRC.

The distribution of response-speed heterogeneity among groups of patients with common clinical factors representing extent of metastases (e.g. number of metastatic sites, number of metastatic lesions), tumor burden (e.g. sum size of target lesions), and response to therapy was consistent with and reinforced previous clinical hypotheses and observations in a quantitative manner. Our observation on the significant association between higher response-speed heterogeneity and shorter survival was very robust for the following three reasons. First, in the multivariable survival analysis, we have adjusted most of the common prognostic factors that were available from our datasets, especially response speed, intuitively considered as the most significant prognostic factor in patients with mCRC and other solid tumors. Second, we have performed multiple sensitivity analyses including leaving out a variable at a time from the multivariable regression models and also constructing response-speed heterogeneity parameter from target lesions in the liver only instead of all target lesions. All these sensitivity analyses provided comparable results. Third, we have externally validated the findings in not only a similar patient cohort who received a different treatment but also a cohort with slightly different baseline patient characteristics, follow-up time, and study design.

Our study has several advantages over some of the previous studies. First, we have utilized individual patient data from previous clinical trials with large sample sizes. Procedures and precautions have been implemented in data collection in clinical trials for quality control. In addition, the prospectively collected patient data for the most part minimize selection bias in this study. Second, the response-speed heterogeneity parameter derived from radiographic data in this study is conceptually plausible and mathematically straightforward, which will likely facilitate the potential application in the clinical setting. Third, we derived the response-speed heterogeneity from a relatively homogenous patient cohort and then validated it in two other patient cohorts. This approach has advantages over some of the previous retrospective studies or studies pooling heterogeneous patient populations to reduce potential confounders and lack of generalizability. Last but not least, from a methodology perspective, we utilized relative size change instead of absolute size change to minimize the effect of the deviation of the data from normal distribution. Furthermore, we derived response-speed heterogeneity from target lesion measurement clearly defined by RECIST criteria instead of inclusion of non-target lesions to reduce the subjectivity at the time of measuring small lesions.^[13-17]^

Our study has several limitations. First and foremost, the data we used were prior to the era of routine molecular profiling in patients with mCRC. The derivation of response-speed heterogeneity may not be affected by molecular alterations of the tumor. Yet, significant prognostic markers such as *RAS/RAF* mutations, MMR/MSI status or CMS subtypes were not made available for us to use in the study, thus, cannot be adjusted in the multivariable models or used for patient stratification. The prognostic role of response-speed heterogeneity in patients with different molecular subgroups should be performed in future studies. Second, the first-line treatments received by the patients included in our study were slightly different from those in current clinical practice. *RAS* mutation information was not used at that time to select patients for panitumumab + chemotherapy (e.g., *RAS* status was not required at entry for the Amgen trial, and the testing was performed later. Patients, *RAS* status was not provided in the Project Data Sphere).^[22]^ Response-speed heterogeneity should be evaluated further with modern patient populations including those who receive other first-line therapies for mCRC and those with ECOG 2, who were otherwise often excluded from clinical trials. Third, in this study, we focused on the development and evaluation of response-speed heterogeneity in patients with previously untreated mCRC. Future direction should extend the generalizability assessment of this response-speed heterogeneity parameter to patients receiving different first-line or later lines of therapy for not only mCRC but also other types of metastatic solid tumors.

## Conclusion

In summary, we established a response-speed heterogeneity parameter in patients with mCRC. It was an independent prognostic factor associated with early disease progression and shorter survival. Complementary to existing molecular and radiographic tumor heterogeneity parameters, it may help practicing oncologists describe tumor response disparity and serve as a new prognostic factor for patients with mCRC.

## Data Availability

All data produced in the present study are available upon reasonable request to the authors.

## Figure Legends

**Online-only Supplemental Figure S1**. CONSORT diagram.

**Online-only Supplemental Figure S2**. Individual tumor response to therapy in representative patients. Dots of the same color connected by solid lines refer to size change over time of an individual target lesion, or sum size (SS) of target lesions. The best response of each target lesion was indicated by “X”. The projection of “X” on the x-axis is t_L_ (for individual lesion) or T_L_ (for sum size). The projection of “X” on the y-axis is LS_L_ or SS_L_. Vertical dashed line indicated the time of disease progression according to RECIST. A lesion,s average relative-size change speed was calculated by “((size at the best response – baseline size) / baseline size) / days on treatment” (See “Patients and Methods” section for other mathematical equations). This parameter estimates a lesion,s response speed to treatment, and can be negative or positive. A negative response speed indicates that a lesion decreases in size after treatment. A positive response speed indicates that treatment was never able to suppress lesion growth. CR: complete response; PR: partial response; SD: stable disease. (a) A patient had PR. Disease progression on day 357 determined by increased sum size of target lesions (non-target lesions had SD; no appearance of new lesions). Response-speed heterogeneity: 1.89×10^−3^ day^-1^. (b) A patient had SD. Disease progression on day 212 determined by increased sum size of both target and non-target lesions (no appearance of new lesions). Response-speed heterogeneity: 2.97×10^−3^ day^-1^. (c) A patient had SD. Disease progression on day 101 determined by increased sum size of target lesions (no non-target lesion; no appearance of new lesions). Response-speed heterogeneity: 2.57×10^−3^ day^-1^. (d) A patient had CR. Disease progression on day 272 determined by appearance of new lesions (sum size of target lesions had CR. non-target lesions had CR). Response-speed heterogeneity: 0 day^-1^.

**Online-only Supplemental Figure S3**. Distributions of response-speed heterogeneity stratified by (a) sex, (b) ethnicity, (c) primary tumor location (colon and rectosigmoid vs. rectal and other), (d) ECOG, (e) age, (f) presence of liver metastasis, (g) presence of lung metastasis, (h) presence of peritoneum metastasis, (i) albumin, and (j) carcinoembryonic antigen in the Amgen (FOLFOX) cohort. *P*-value from Wilcoxon rank-sum test for comparing two groups, or from Kruskal-Wallis test for comparing three or more groups.

