## Supplemental Figures for "Tumor response-speed heterogeneity as a novel prognostic factor in patients with mCRC"

**Supplemental Figure S1.** CONSORT diagram.

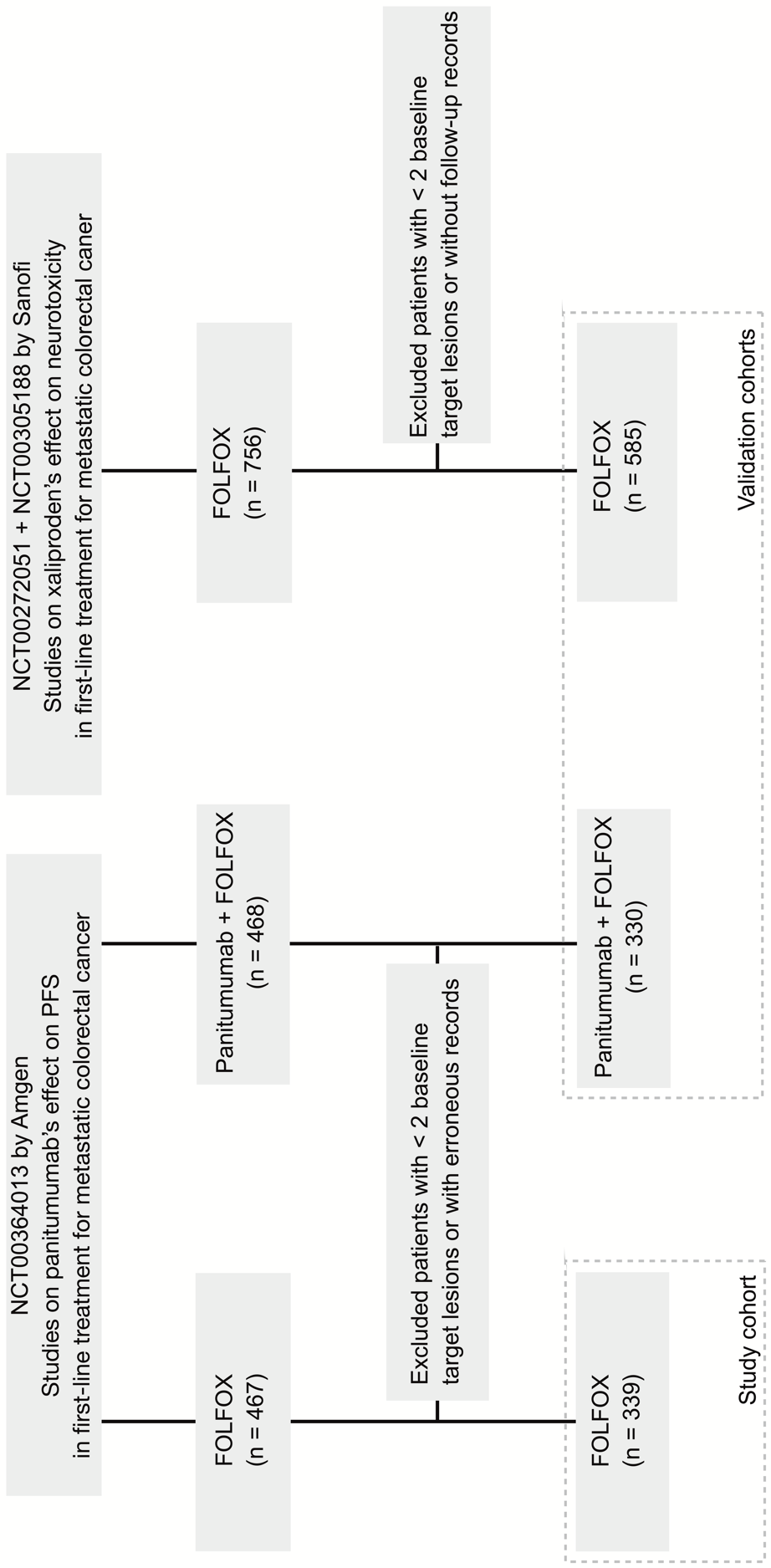

**Supplemental Figure S2.** Individual tumor response to therapy in representative patients. Dots of the same color connected by solid lines refer to size change over time of an individual target lesion, or sum size (SS) of target lesions. The best response of each target lesion was indicated by “X”. The projection of “X” on the x-axis is t_L_ (for individual lesion) or T_L_ (for sum size). The projection of “X” on the y-axis is LS_L_ or SS_L_. Vertical dashed line indicated the time of disease progression according to RECIST. A lesion’s average relative-size change speed was calculated by “( (size at the best response – baseline size) / baseline size) / days on treatment” (See “Patients and Methods” section for other mathematical equations). This parameter estimates a lesion’s response speed to treatment, and can be negative or positive. A negative response speed indicates that a lesion decreases in size after treatment. A positive response speed indicates that treatment was never able to suppress lesion growth. CR: complete response; PR: partial response; SD: stable disease. (a) A patient had PR. Disease progression on day 357 determined by increased sum size of target lesions (non-target lesions had SD; no appearance of new lesions). Response-speed heterogeneity: 1.89×10^-3^ day^-1^. (b) A patient had SD. Disease progression on day 212 determined by increased sum size of both target and non-target lesions (no appearance of new lesions). Response-speed heterogeneity: 2.97×10^-3^ day^-1^. (c) A patient had SD. Disease progression on day 101 determined by increased sum size of target lesions (no non-target lesion; no appearance of new lesions). Response-speed heterogeneity: 2.57×10^-3^ day^-1^. (d) A patient had CR. Disease progression on day 272 determined by appearance of new lesions (sum size of target lesions had CR. non-target lesions had CR). Response-speed heterogeneity: 0 day^-1^.

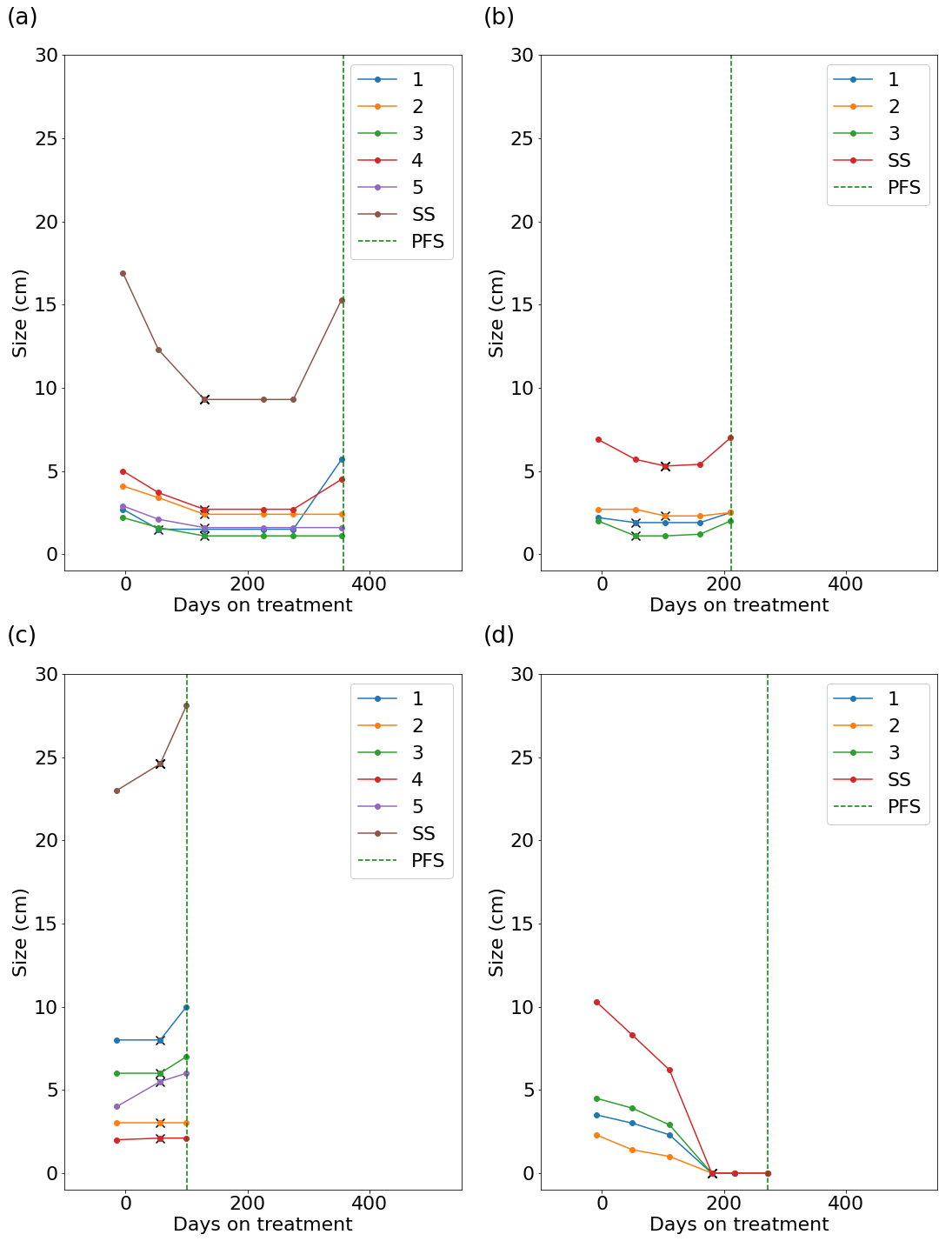

**Supplemental Figure S3**. Distributions of response-speed heterogeneity stratified by (a) sex, (b) ethnicity, (c) primary tumor location (colon and rectosigmoid vs. rectal and other), (d) ECOG, (e) age, (f) presence of liver metastasis, (g) presence of lung metastasis, (h) presence of peritoneum metastasis, (i) albumin, and (j) carcinoembryonic antigen in the Amgen (FOLFOX) cohort. *P*-value from Wilcoxon rank-sum test for comparing two groups, or from Kruskal-Wallis test for comparing three or more groups.

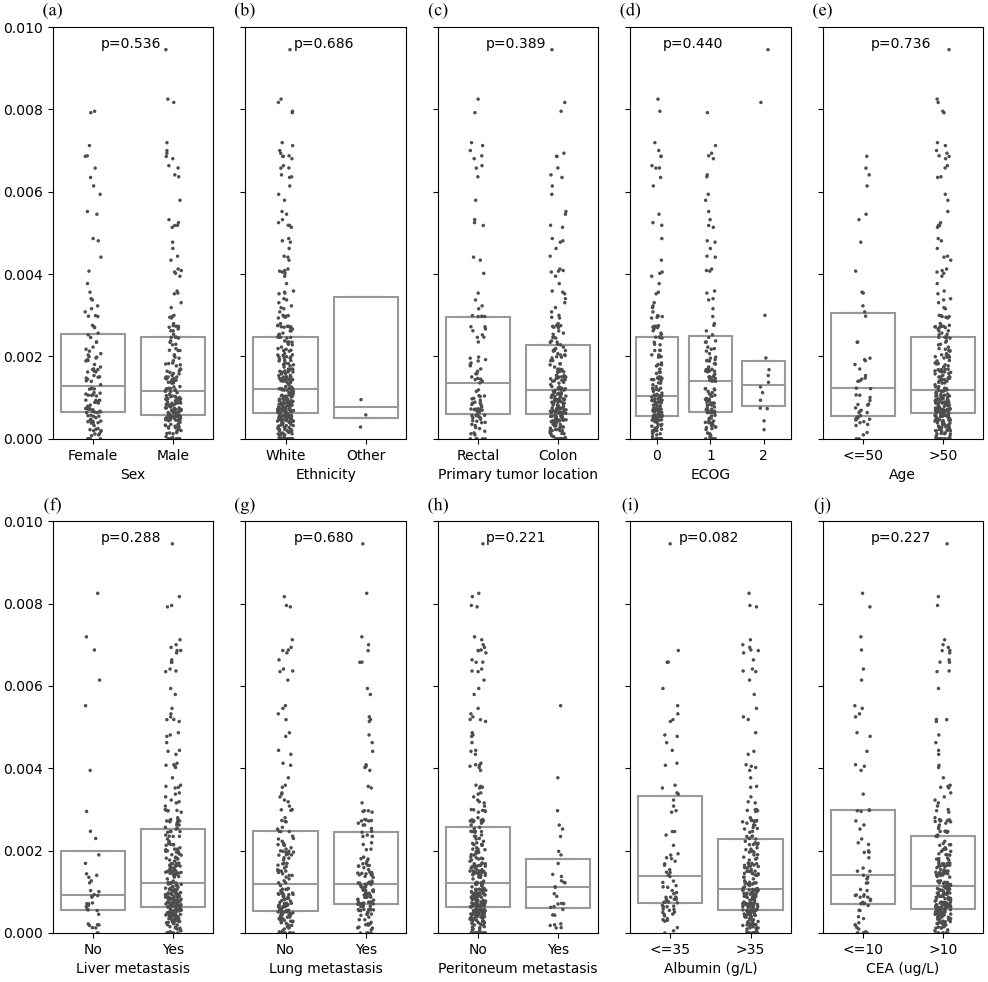
