## Supplemental Tables for "Tumor response-speed heterogeneity as a novel prognostic factor in patients with mCRC"

**Supplemental Table S1.** Univariate and multivariable analyses of PFS for the Amgen (FOLFOX) cohort’s subgroup: response-speed heterogeneity was only based on liver target lesions.

| Variable | Univariate analysis | | | | Multivariable analysis | | | |
| --- | --- | --- | --- | --- | --- | --- | --- | --- |
|  | HR | 95% CI |  | *P* | HR | 95% CI |  | *P* |
| Liver response-speed heterogeneity (every 0.01 day^-1^ increase) | 3.42 | 1.94 | - 6.02 | <0.0001 | 5.31 | 2.83 | - 9.95 | <0.0001 |
| Liver baseline size heterogeneity (every 0.01 increase) | 1.001 | 0.99 | - 1.01 | 0.77 | 0.99 | 0.98 | - 1.003 | 0.16 |
| Sum size of target lesions (every 1 cm increase) | 1.02 | 1.01 | - 1.04 | 0.00069 | 1.01 | 0.99 | - 1.03 | 0.27 |
| Response speed (every 0.01 day^-1^ increase) | 0.30 | 0.15 | - 0.62 | 0.0011 | 0.24 | 0.12 | - 0.48 | <0.0001 |
| Age (every 10 years increase) | 0.94 | 0.82 | - 1.08 | 0.37 | 0.83 | 0.72 | - 0.96 | 0.014 |
| Albumin (every 0.1 g/dL increase) | 0.96 | 0.94 | - 0.99 | 0.0048 | 0.97 | 0.94 | - 0.996 | 0.026 |
| CEA (every 10 ug/L increase) | 1.001 | 0.9998 | - 1.002 | 0.15 | 0.9999 | 0.999 | - 1.001 | 0.87 |
| Male (vs. female) | 0.99 | 0.76 | - 1.28 | 0.92 | 0.997 | 0.74 | - 1.33 | 0.98 |
| Lung metastasis | 1.32 | 1.01 | - 1.71 | 0.040 | 1.26 | 0.94 | - 1.68 | 0.12 |
| Peritoneum metastasis | 1.45 | 0.89 | - 2.36 | 0.13 | 1.33 | 0.78 | - 2.22 | 0.27 |
| Lymph node metastasis | 1.28 | 0.98 | - 1.68 | 0.071 | 0.99 | 0.74 | - 1.34 | 0.97 |
| Number of lesions >=5 (vs. 2-4) | 1.36 | 0.99 | - 1.87 | 0.055 | 1.01 | 0.69 | - 1.48 | 0.95 |
| ECOG 1 (vs. 0) | 1.60 | 1.22 | - 2.10 | 0.00069 | 1.37 | 1.02 | - 1.83 | 0.037 |
| ECOG 2 (vs. 0) | 2.91 | 1.62 | - 5.23 | 0.00035 | 2.02 | 0.99 | - 4.14 | 0.054 |

**Supplemental Table S2.** Univariate analysis of PFS for the Sanofi (FOLFOX) cohort.

| Variable | Univariate analysis | | | |
| --- | --- | --- | --- | --- |
|  | HR | 95% CI | | *P* |
| Response-speed heterogeneity (every 0.01 day^-1^ increase) | 1.56 | 1.05 | - 2.31 | 0.027 |
| Baseline size heterogeneity (every 0.01 increase) | 1.004 | 0.9999 | - 1.01 | 0.054 |
| Sum size of target lesions (every 1 cm increase) | 1.02 | 1.01 | - 1.03 | <0.0001 |
| Response speed (every 0.01 day^-1^ increase) | 0.41 | 0.28 | - 0.62 | <0.0001 |
| Age (every 10 years increase) | 0.93 | 0.86 | - 1.01 | 0.070 |
| CEA (every 10 ug/L increase) | 0.9999 | 0.999 | - 1.001 | 0.81 |
| Male (vs. female) | 0.87 | 0.73 | - 1.03 | 0.11 |
| Liver metastasis | 1.24 | 0.98 | - 1.58 | 0.077 |
| Lung metastasis | 1.34 | 1.13 | - 1.60 | 0.00089 |
| Peritoneum metastasis | 1.22 | 0.87 | - 1.70 | 0.25 |
| Lymph node metastasis | 1.06 | 0.89 | - 1.27 | 0.52 |
| Number of lesions >=5 (vs. 2-4) | 1.21 | 1.02 | - 1.44 | 0.028 |
| ECOG 1 (vs. 0) | 1.16 | 0.97 | - 1.39 | 0.10 |
| ECOG 2 (vs. 0) | 2.13 | 1.22 | - 3.72 | 0.0077 |

**Supplemental Table S3.** Univariate analysis of PFS for the Sanofi (panitumumab + FOLFOX) cohort.

| Variable | Univariate analysis | | | |
| --- | --- | --- | --- | --- |
|  | HR | 95% CI | | *P* |
| Response-speed heterogeneity (every 0.01 day^-1^ increase) | 2.09 | 1.27 | - 3.42 | 0.0037 |
| Baseline size heterogeneity (every 0.01 increase) | 1.001 | 0.996 | - 1.01 | 0.64 |
| Sum size of target lesions (every 1 cm increase) | 1.01 | 0.99 | - 1.02 | 0.32 |
| Response speed (every 0.01 day^-1^ increase) | 0.74 | 0.43 | - 1.28 | 0.28 |
| Age (every 10 years increase) | 1.08 | 0.96 | - 1.22 | 0.21 |
| Albumin (every 0.1 g/dL increase) | 0.97 | 0.95 | - 0.99 | 0.0015 |
| CEA (every 10 ug/L increase) | 1.0002 | 0.9999 | - 1.001 | 0.19 |
| Male (vs. female) | 0.94 | 0.74 | - 1.19 | 0.60 |
| Liver metastasis | 0.89 | 0.62 | - 1.29 | 0.55 |
| Lung metastasis | 0.95 | 0.76 | - 1.19 | 0.67 |
| Peritoneum metastasis | 1.49 | 1.02 | - 2.18 | 0.037 |
| Lymph node metastasis | 1.01 | 0.80 | - 1.27 | 0.94 |
| Number of lesions >=5 (vs. 2-4) | 1.30 | 1.004 | - 1.68 | 0.047 |
| ECOG 1 (vs. 0) | 1.33 | 1.05 | - 1.68 | 0.016 |
| ECOG 2 (vs. 0) | 3.23 | 2.03 | - 5.13 | <0.0001 |

**Supplemental Table S4.** Univariate and multivariable analyses of OS for the Amgen (FOLFOX) cohort.

| Variable | Univariate analysis | | | | Multivariable analysis | | | |
| --- | --- | --- | --- | --- | --- | --- | --- | --- |
|  | HR | 95% CI |  | *P* | HR | 95% CI |  | *P* |
| Response-speed heterogeneity (every 0.01 day^-1^ increase) | 3.21 | 2.09 | - 4.93 | <0.0001 | 2.57 | 1.64 | - 4.01 | <0.0001 |
| Baseline size heterogeneity (every 0.01 increase) | 1.01 | 1.0003 | - 1.01 | 0.041 | 0.99 | 0.99 | - 1.003 | 0.24 |
| Sum size of target lesions (every 1 cm increase) | 1.05 | 1.03 | - 1.06 | <0.0001 | 1.04 | 1.02 | - 1.06 | 0.00010 |
| Response speed (every 0.01 day^-1^ increase) | 0.49 | 0.29 | - 0.84 | 0.0093 | 0.49 | 0.29 | - 0.84 | 0.0092 |
| Age (every 10 years increase) | 1.09 | 0.96 | - 1.24 | 0.17 | 0.998 | 0.87 | - 1.14 | 0.98 |
| Albumin (every 0.1 g/dL increase) | 0.92 | 0.90 | - 0.94 | <0.0001 | 0.94 | 0.91 | - 0.97 | <0.0001 |
| CEA (every 10 ug/L increase) | 1.0001 | 0.9999 | - 1.0003 | 0.52 | 0.99997 | 0.9997 | - 1.0002 | 0.84 |
| Male (vs. female) | 1.07 | 0.83 | - 1.37 | 0.61 | 1.22 | 0.91 | - 1.63 | 0.18 |
| Liver metastasis | 1.67 | 1.12 | - 2.51 | 0.013 | 1.57 | 0.97 | - 2.55 | 0.064 |
| Lung metastasis | 1.24 | 0.98 | - 1.58 | 0.074 | 1.16 | 0.88 | - 1.53 | 0.29 |
| Peritoneum metastasis | 0.97 | 0.64 | - 1.45 | 0.87 | 0.91 | 0.58 | - 1.45 | 0.70 |
| Lymph node metastasis | 1.52 | 1.19 | - 1.95 | 0.00077 | 1.37 | 1.04 | - 1.81 | 0.027 |
| Number of lesions >=5 (vs. 2-4) | 1.60 | 1.20 | - 2.14 | 0.0014 | 0.92 | 0.64 | - 1.33 | 0.67 |
| ECOG 1 (vs. 0) | 1.65 | 1.28 | - 2.11 | <0.0001 | 1.57 | 1.19 | - 2.07 | 0.0015 |
| ECOG 2 (vs. 0) | 3.84 | 2.19 | - 6.74 | <0.0001 | 1.48 | 0.76 | - 2.87 | 0.25 |

**Supplemental Table S5.** Univariate and multivariable analyses of OS for the Sanofi (FOLFOX) cohort.

| Variable | Univariate analysis | | | | Multivariable analysis | | | |
| --- | --- | --- | --- | --- | --- | --- | --- | --- |
|  | HR | 95% CI |  | *P* | HR | 95% CI |  | *P* |
| Response-speed heterogeneity (every 0.01 day^-1^ increase) | 1.99 | 1.20 | - 3.31 | 0.0078 | 1.55 | 0.91 | - 2.66 | 0.11 |
| Baseline size heterogeneity (every 0.01 increase) | 1.01 | 1.002 | - 1.01 | 0.0037 | 1.002 | 0.996 | - 1.01 | 0.45 |
| Sum size of target lesions (every 1 cm increase) | 1.04 | 1.03 | - 1.05 | <0.0001 | 1.03 | 1.02 | - 1.05 | <0.0001 |
| Response speed (every 0.01 day^-1^ increase) | 0.25 | 0.17 | - 0.38 | <0.0001 | 0.23 | 0.15 | - 0.36 | <0.0001 |
| Age (every 10 years increase) | 0.85 | 0.77 | - 0.94 | 0.0015 | 0.90 | 0.81 | - 1.01 | 0.062 |
| CEA (every 10 ug/L increase) | 1.0001 | 0.999 | - 1.001 | 0.78 | 0.9996 | 0.998 | - 1.001 | 0.44 |
| Male (vs. female) | 0.80 | 0.63 | - 1.003 | 0.053 | 0.70 | 0.55 | - 0.91 | 0.0069 |
| Liver metastasis | 1.58 | 1.13 | - 2.22 | 0.0076 | 1.95 | 1.29 | - 2.95 | 0.0014 |
| Lung metastasis | 1.10 | 0.88 | - 1.39 | 0.41 | 1.32 | 1.02 | - 1.72 | 0.036 |
| Peritoneum metastasis | 1.84 | 1.27 | - 2.67 | 0.0014 | 1.87 | 1.23 | - 2.84 | 0.0035 |
| Lymph node metastasis | 1.29 | 1.02 | - 1.62 | 0.035 | 1.23 | 0.94 | - 1.60 | 0.13 |
| Number of lesions >=5 (vs. 2-4) | 1.32 | 1.05 | - 1.66 | 0.016 | 0.81 | 0.60 | - 1.10 | 0.18 |
| ECOG 1 (vs. 0) | 1.57 | 1.24 | - 1.98 | 0.00017 | 1.41 | 1.09 | - 1.82 | 0.0083 |
| ECOG 2 (vs. 0) | 3.94 | 2.13 | - 7.23 | <0.0001 | 2.63 | 1.25 | - 5.54 | 0.011 |

**Supplemental Table S6.** Univariate and multivariable analyses of OS for the Amgen (panitumumab + FOLFOX) cohort.

| Variable | Univariate analysis | | | | Multivariable analysis | | | |
| --- | --- | --- | --- | --- | --- | --- | --- | --- |
|  | HR | 95% CI |  | *P* | HR | 95% CI |  | *P* |
| Response-speed heterogeneity (every 0.01 day^-1^ increase) | 1.83 | 1.06 | - 3.18 | 0.033 | 1.88 | 1.04 | - 3.39 | 0.037 |
| Baseline size heterogeneity (every 0.01 increase) | 1.01 | 1.001 | - 1.01 | 0.015 | 1.004 | 0.996 | - 1.01 | 0.30 |
| Sum size of target lesions (every 1 cm increase) | 1.02 | 1.01 | - 1.04 | <0.0001 | 1.01 | 0.99 | - 1.03 | 0.28 |
| Response speed (every 0.01 day^-1^ increase) | 0.46 | 0.31 | - 0.68 | 0.00012 | 0.41 | 0.28 | - 0.62 | <0.0001 |
| Age (every 10 years increase) | 1.03 | 0.90 | - 1.18 | 0.65 | 1.05 | 0.90 | - 1.23 | 0.51 |
| Albumin (every 0.1 g/dL increase) | 0.95 | 0.93 | - 0.97 | <0.0001 | 0.97 | 0.94 | - 0.995 | 0.021 |
| CEA (every 10 ug/L increase) | 1.0002 | 0.999996 | - 1.0004 | 0.054 | 1.0002 | 0.9999 | - 1.0005 | 0.12 |
| Male (vs. female) | 0.97 | 0.75 | - 1.26 | 0.83 | 0.93 | 0.70 | - 1.23 | 0.61 |
| Liver metastasis | 1.10 | 0.73 | - 1.64 | 0.66 | 0.74 | 0.46 | - 1.20 | 0.22 |
| Lung metastasis | 1.16 | 0.91 | - 1.48 | 0.24 | 1.12 | 0.84 | - 1.48 | 0.44 |
| Peritoneum metastasis | 1.39 | 0.93 | - 2.07 | 0.11 | 0.93 | 0.58 | - 1.47 | 0.75 |
| Lymph node metastasis | 1.04 | 0.81 | - 1.33 | 0.78 | 0.98 | 0.74 | - 1.29 | 0.88 |
| Number of lesions >=5 (vs. 2-4) | 1.58 | 1.18 | - 2.13 | 0.0023 | 1.21 | 0.85 | - 1.72 | 0.29 |
| ECOG 1 (vs. 0) | 1.61 | 1.24 | - 2.08 | 0.00028 | 1.42 | 1.06 | - 1.90 | 0.020 |
| ECOG 2 (vs. 0) | 3.76 | 2.33 | - 6.06 | <0.0001 | 2.16 | 1.12 | - 4.16 | 0.022 |
